# How are combinations of physical activity, sedentary behaviour and sleep related to cognitive function in older adults? A systematic review

**DOI:** 10.1101/2021.09.07.21263199

**Authors:** Maddison L Mellow, Alyson J Crozier, Dorothea Dumuid, Alexandra T Wade, Mitchell R Goldsworthy, Jillian Dorrian, Ashleigh E Smith

**Author notes:** Corresponding author: Dr Ashleigh E. Smith. Alliance for Research in Exercise, Nutrition and Activity, Allied Health and Human Performance, University of South Australia, GPO Box 2471, Adelaide, South Australia 5001.

## Abstract

The relationships between cognitive function and each of physical activity, sleep and sedentary behaviour in older adults are well documented. However, these three “time use” behaviours are co-dependent parts of the 24-hour day (spending time in one leaves less time for the others), and their best balance for cognitive function in older adults is still largely unknown. This systematic review summarises the existing evidence on the associations between combinations of two or more time-use behaviours and cognitive function in older adults. Embase, Pubmed, PsycInfo, Medline and Emcare databases were searched in March 2020 and updated in May 2021, returning a total of 25,289 papers for screening. A total of 23 studies were included in the synthesis, spanning >23,000 participants (mean age 71 years). Findings support previous evidence that spending more time in physical activity and limiting sedentary behaviour is broadly associated with better cognitive outcomes in older adults. Higher proportions of moderate-vigorous physical activity in the day were most frequently associated with better cognitive function. Some evidence suggests that certain types of sedentary behaviour may be positively associated with cognitive function, such as reading or computer use. Sleep duration appears to share an inverted U-shaped relationship with cognition, as too much or too little sleep is negatively associated with cognitive function. This review highlights considerable heterogeneity in methodological and statistical approaches, and encourages a more standardised, transparent approach to capturing important daily behaviours in older adults. Investigating all three time-use behaviours together against cognitive function using suitable statistical methodology is strongly recommended to further our understanding of optimal 24-hour time-use for brain function in aging.

## 1. Introduction

The benefits of engaging in sufficient physical activity (PA), limiting sedentary behaviour (SB) and maintaining healthy sleep patterns extend beyond physical health to brain health. A recent report by Livingston et al. (2020) identified physical inactivity in late life as a modifiable risk factor for dementia, which is currently the 7^th^ leading cause of death worldwide (World Health Organisation, 2020). Poor sleep was also identified as a modifiable risk factor for dementia, although evidence has been more sparse and generally inconclusive (Livingston et al., 2020). In the absence of a pharmacological cure for dementia, the focus of research has now largely turned to prevention, and understanding how modifiable risk factors such as PA, SB and sleep are related to cognitive function and future dementia risk in older adulthood.

The relationships of PA, SB and sleep behaviours with cognitive function in older adults are well documented in the literature. Spending sufficient time engaging in PA is widely recognized as a beneficial health behaviour for maintaining or improving cognitive function in older adulthood (Erickson et al., 2019), whilst spending excessive time in SB (e.g. sitting, watching television) is often negatively associated with cognitive function (Coelho et al., 2020). Findings of a recent systematic review demonstrated that greater amounts of device-measured total PA or moderate-vigorous PA (MVPA) were positively associated with cognitive function in older adults, whilst greater amounts of objectively measured SB was negatively associated with cognitive function (Rojer et al., 2021). In addition, an important relationship between sleep and cognitive function has been reported in the literature. Poorer self-reported sleep quality in older adults (aged 65+ years) has been negatively associated with cognitive function, whilst sleep duration appears to share an inverted U-shaped relationship with cognition, as achieving above or below the typical duration (7-9 hours) has been associated with poorer cognitive function (Nebes et al., 2009; Yaffe et al., 2014). Taken together, there is sufficient evidence to suggest that PA, SB and sleep affect brain health and function in later life.

One major limitation of previous research is that most studies have investigated PA, SB or sleep as stand-alone behaviours in separate analytical models. However, these three behaviours are mutually exclusive and exhaustive components of a 24-hour day (Figure 1). Any increase in time spent in one behaviour must result in an equal and opposite change in at least one other behaviour. Studies which have previously investigated the relationship between only one time-use behaviour and cognitive outcomes are therefore unable to disentangle the influence of one time-use behaviour from the others. Indeed, emerging evidence suggests that excessive time spent in SB may ‘reverse’ the positive effects of PA on brain health and function (Voss et al., 2014). Similarly, it is well understood that PA and sleep are strongly related, as increasing PA may improve sleep quality, whilst poor sleep quality may reduce engagement in PA (Best et al., 2019; Wang and Boros, 2021). It is critical to consider combinations of time-use behaviours to gain a holistic understanding of the ‘best day’ for cognitive function. One approach which aims to overcome this limitation considers how combinations of time-use behaviours within the 24-hour day, or *time-use compositions*, are associated with cognitive function. This approach has gained traction in the past decade, and has been used to understand the best combination of all three time-use behaviours in the 24-hour day for outcomes including brain plasticity (Smith et al., 2021), adiposity (Dumuid et al., 2021) and skeletal health (Dumuid et al., 2020). Understanding the optimal combinations of PA, SB and sleep for cognitive function in older adults is a key step to informing 24-hour movement guidelines for dementia prevention.

**Figure 1:**
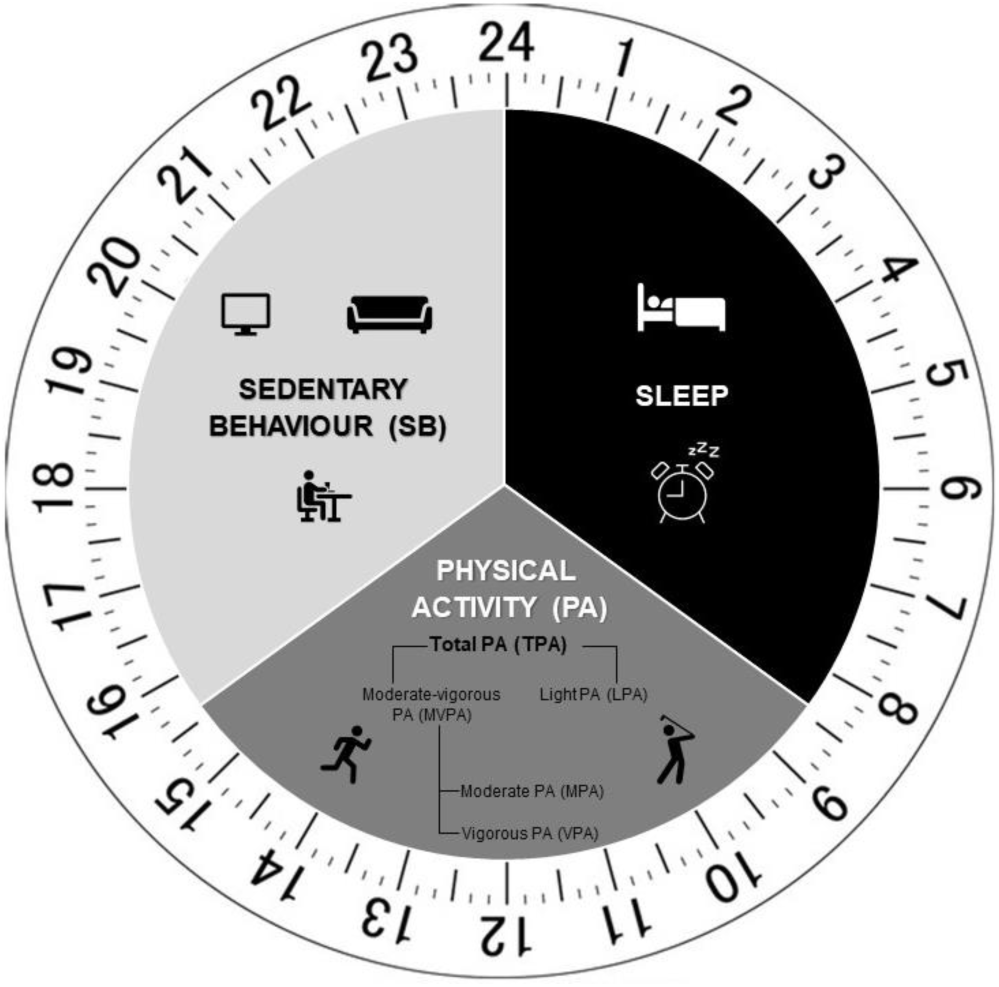
Time-use varies across the 24-hour day.

Our previous narrative review explored the separate relationships between PA, sleep and SB with brain structure, function and neurodegenerative disease risk across the lifespan (Mellow et al., 2019). However, to our knowledge, the current evidence surrounding combinations of all three time-use behaviours (PA, SB and sleep) and cognitive function across several cognitive domains in older adults is yet to be synthesised. Therefore, the current systematic review summarises the existing evidence of how combinations of PA, sleep and SB are associated with cognitive function in older adults.

## 2. Methods

This systematic review was conducted in accordance with the Preferred Reporting Items for Systematic Reviews and Meta-Analyses (PRISMA) statement (Page et al., 2021) and was prospectively registered with PROSPERO (registration ID: CRD42020184511).

### 2.1 Inclusion criteria

We included cross-sectional observational studies, and intervention or longitudinal studies if sufficient baseline data could be extracted. Studies were included if: the population had a mean age of ≥60 years; the study included at least one healthy control group (i.e., with no psychological or neurological diagnoses for comparison to clinical sub-groups where applicable); the study measured and reported on any combination of two or more time-use behaviours (PA, sleep or SB) as independent variables (Saunders et al., 2016). Studies which only investigated one time-use behaviour, or only included the second time-use behaviour as a covariate in the model, were excluded. Measures of time-use behaviours could be objective (e.g. using accelerometers, pedometers, or polysomnography) or subjective (e.g. self-report questionnaires or time use diaries). Included studies also needed to report a measure of cognitive function as the dependent variable. To be eligible, measures of cognitive function were required to be valid and reliable measures of global cognitive function (e.g. Mini Mental State Examination (MMSE) (Folstein et al., 1983) or Montreal Cognitive Assessment (MoCA) (Nasreddine et al., 2005)) or specific cognitive domains (e.g. computer or paper-and-pencil based measures of executive function, memory, or processing speed). Measures of *subjective* cognitive function, such as single questions asking the participant how they were feeling about their cognitive state, were not included if they were a stand-alone cognitive measure (e.g. not used in addition to an objective measure).

### 2.2 Search strategy

The search strategy was developed in consultation with an academic librarian, and used terms related to PA, SB, sleep, and cognitive function (see Appendix A for full list of search terms). Initial searches were conducted on Embase, PubMed, PsycInfo, Medline and Emcare databases on 15^th^ March 2020, and were updated on 26^th^ May 2021. No limits were applied to date of publication, however only studies published in English were included in this review due to lack of resources to translate non-English papers.

### 2.3 Study screening

Studies collated through the database searches were exported to Endnote (X9). Duplicates were removed firstly using the automated ‘remove duplicates’ function, followed by manual screening. Remaining studies were then uploaded to Covidence for screening. Two researchers (MM and AC or AW) independently screened titles and abstracts, and irrelevant studies were excluded. The full text of each paper was then screened for relevance and eligibility by two researchers (MM and AC). Any conflicts that arose were discussed with a separate researcher (AS) and final decisions were reached via consensus. Updated searches were screened by MM at title and abstract level, and full text papers were screened by both MM and AC. MM extracted relevant data to an excel spreadsheet including the following information: author; date of publication; title; study aims; cognitive measure used; PA, SB and/or sleep measure used; analysis approach; main findings; study limitations; study conclusions. These extractions were checked by AC or AW for accuracy.

### 2.4 Data synthesis

Study outcomes were firstly grouped by cognitive domain, whereby cognitive tests used in each study were assigned to seven broad ability domains using the Cattell-Horn-Carroll-Miyake (CHC-M) taxonomy outlined by Webb et al. (2018). Cognitive domains in this taxonomy include global cognition, executive function, processing speed, general short-term memory, visual processing, fluid reasoning, and long-term memory and retrieval (Webb et al., 2018). Meta-analyses were not conducted because of insufficient homogeneity in measures, methods and statistical approaches used. Instead, a narrative synthesis of results was performed for each cognitive domain. Results are discussed in terms of the most optimal combination of time-use behaviours for performance in each cognitive domain, and/or the relationships between time-use behaviours and cognitive outcomes.

### 2.5 Quality and risk of bias assessment

The Office of Health Assessment and Translation (OHAT) Risk of Bias Tool for Human and Animal Studies was used to assess risk of bias and internal validity (US Department of Health and Human Services, 2015). Six questions from the tool relating to cross-sectional research were assessed for each study. Each question required a score which reflected the risk of bias: as per the original tool, ‘++’ reflects *definitely low* risk of bias, ‘+’ reflects *probably low* risk of bias, ‘-’ reflects *probably high* risk of bias, and ‘- -’ (double negative) reflects *definitely high* risk of bias. For ease of comparison, these scores were recorded as +3, +2, +1 and 0 scores respectively, with the highest possible total score of 18 reflecting the lowest risk of bias, and the lowest possible total score of 0 reflecting highest risk of bias. Additionally, ‘NR’ reflects *not reported* and is classified in the same category as *probably high*. MM conducted the risk of bias analysis. A full list of the six items is provided in Appendix B.

## 3. Results

### 3.1 Study demographics

#### 3.1.1 Study and participant demographics

Initial searches of the databases (March 2020) returned 23,408 references, 1,248 of which were removed during duplicate screening. Title and abstract screening of the remaining 22,160 papers resulted in 128 papers remaining in the full text screening phase. One hundred and nine of these studies were excluded at the full text phase (see Figure 2 for list of reasons), and a final 19 articles met inclusion for the narrative synthesis. The updated searches (2021) returned a further 1,881 references, 1,856 of which were excluded at title and abstract level. The full text of 25 studies were screened, with 4 meeting eligibility criteria. Thus, a total of 23 papers were included in the narrative synthesis of this review.

**Figure 2:**
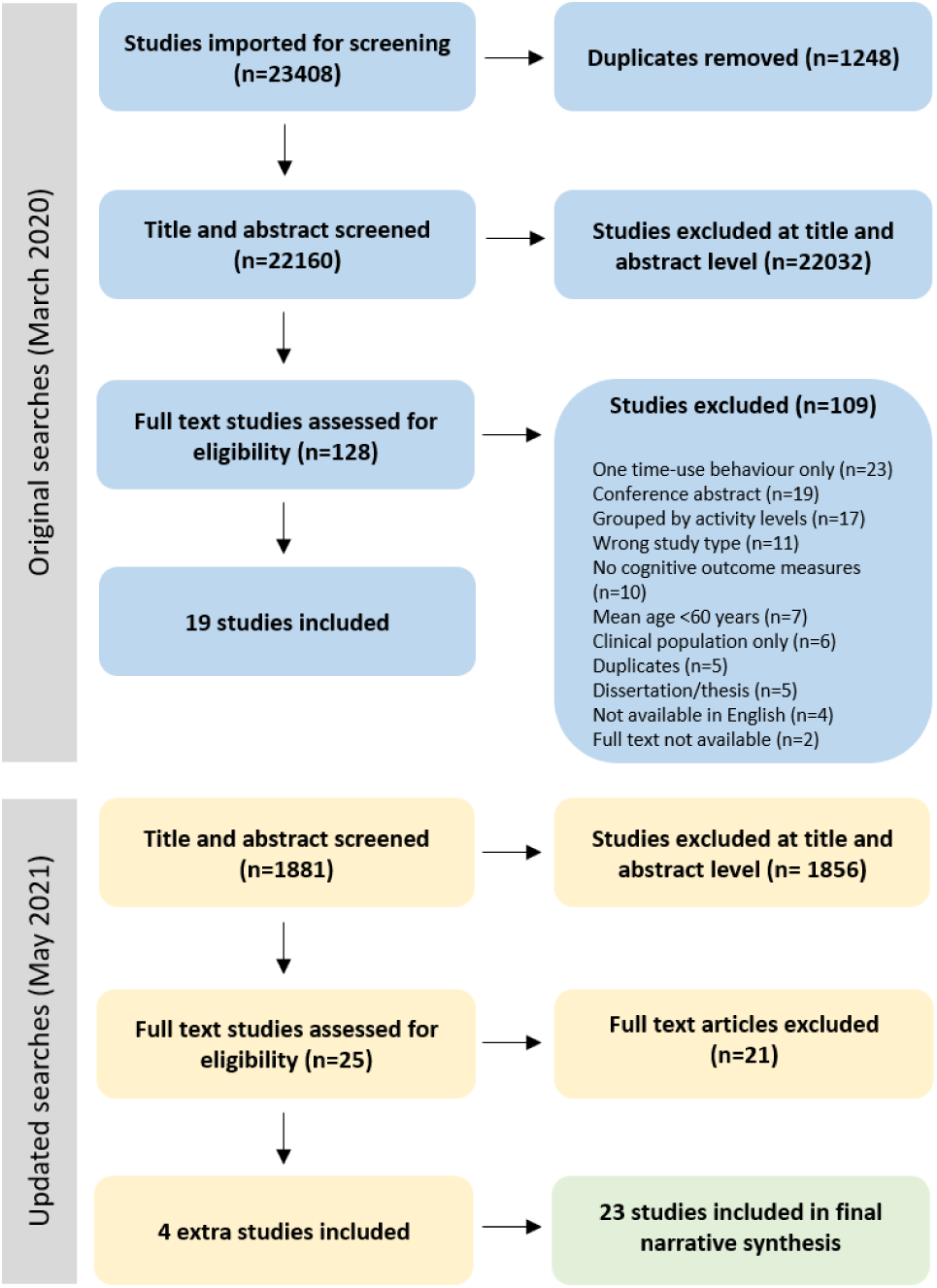
PRISMA flow diagram describing the screening process, including total number of studies (n) included and excluded at each level, and reasons for exclusion.

The 23 studies included in the narrative synthesis spanned 21,829 participants collectively. The average age of participants was 71.3 ± 5.4 years, and 44% of participants were male. Three studies (Amagasa et al., 2019; Falck et al., 2017; Suzuki et al., 2020) included a sub-sample with cognitive decline or probable cognitive decline, based on cut-off scores on cognitive screening tools (<23 on MMSE, <26 on MoCA or <88 on ACE-III, respectively). One study (Anastasiou et al., 2018) included participants with mild cognitive impairment and participants with a diagnosis of dementia, as well as healthy controls. Additionally, Bollaert and Motl (2019) included a sub-sample of older adults diagnosed with Multiple Sclerosis. All 23 studies included a measure of PA, with 18 studies including a second time use measure of SB, and 9 studies including a measure of sleep. Four studies included measures of all three time-use behaviours (Anastasiou et al., 2018; Fanning et al., 2017; Vance et al., 2016; Wei et al., 2021).

Categorisation of cognitive tests into sub-domains outlined in Webb et al. (2018) identified seven cognitive domains (Table 1). Of the 23 included studies, 11 included a measure of global cognition, eight a measure of executive function, 10 a measure of long-term memory and retrieval, 10 a measure of processing speed, three a measure of general short-term memory, two a measure of visual processing and one study included a measure of fluid reasoning. Two studies (Burzynska et al., 2020; Wilckens et al., 2018) did not report results for individual cognitive tests, and instead reported associations between time-use behaviours and pre-determined cognitive domains (composites). These studies were included in the analyses as their classification of cognitive tests into domains matched the categorisation framework used in this review.

**Table 1.**
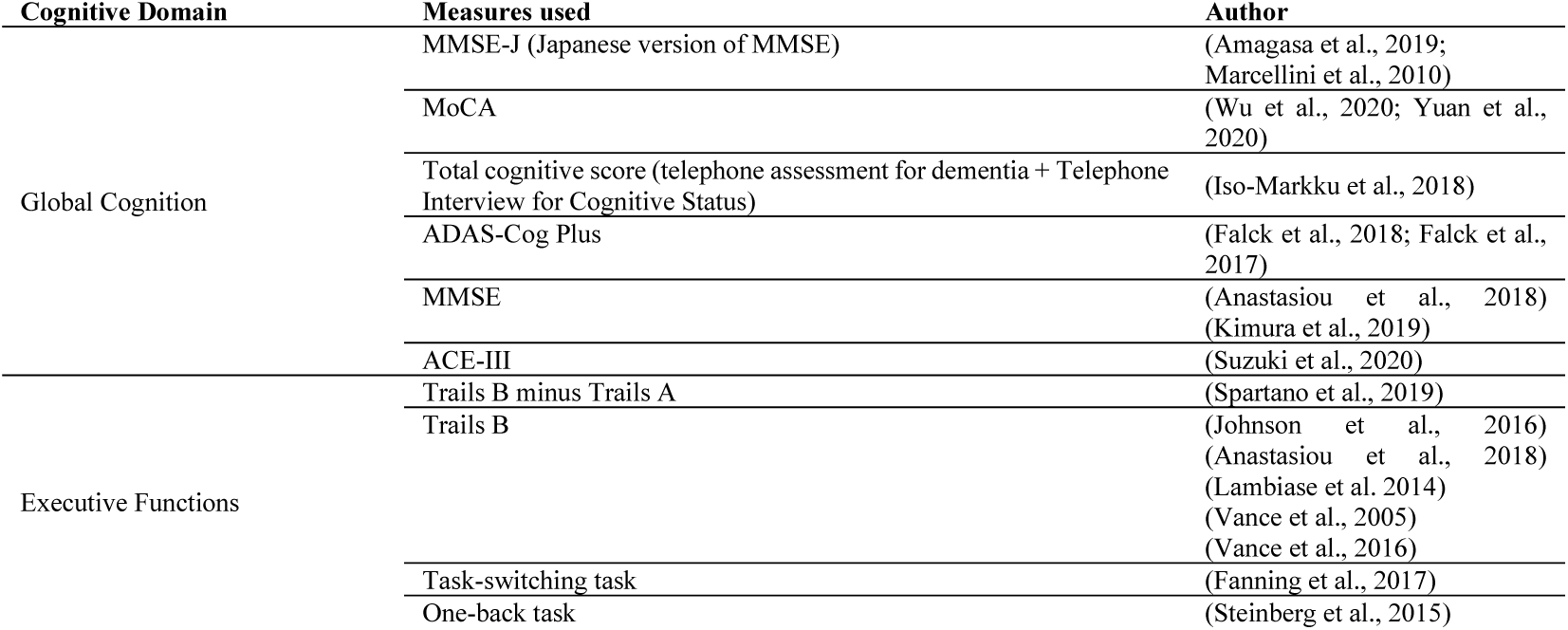
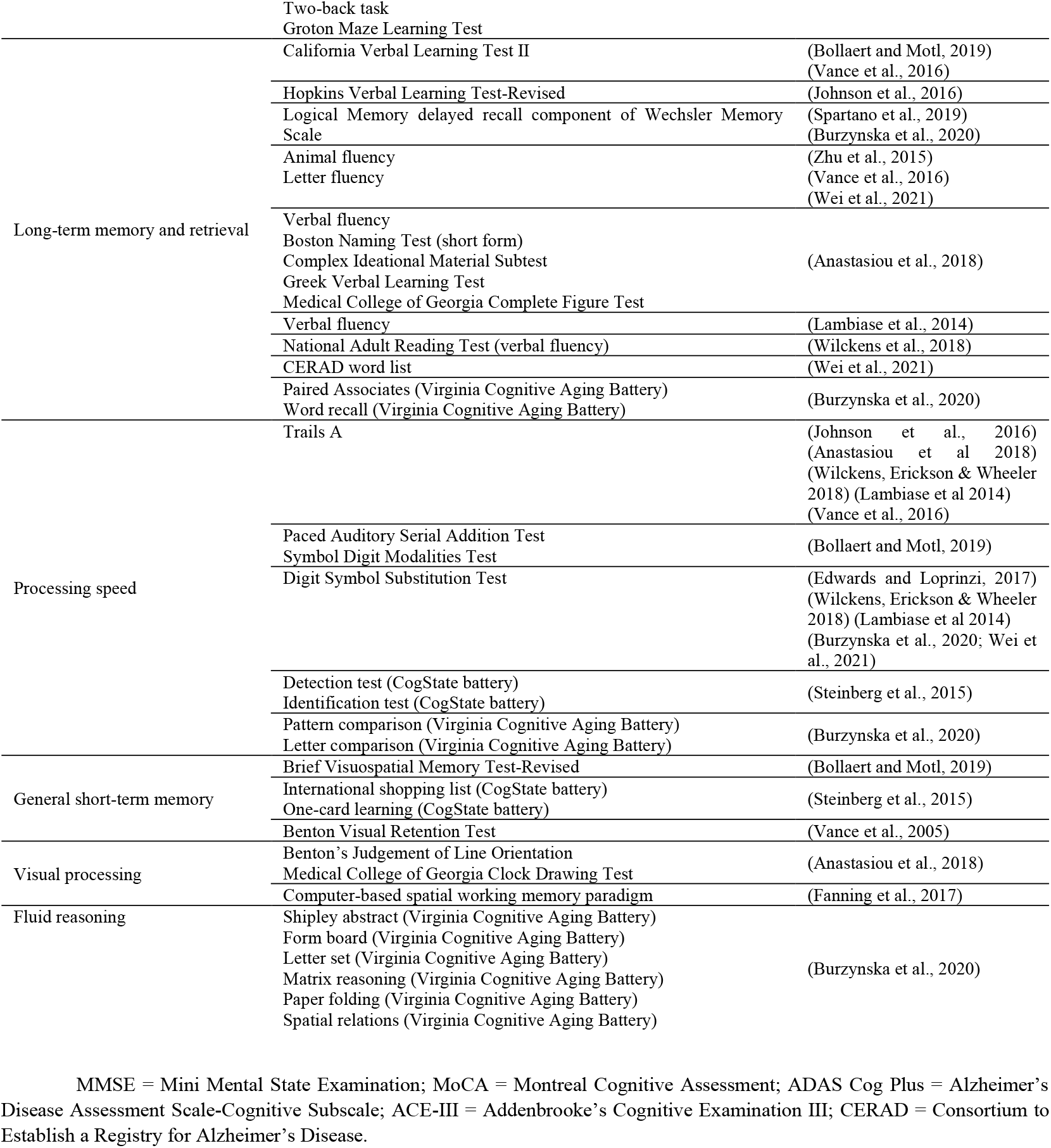
Categorisation of cognitive tasks per cognitive domain.

### 3.2 Measures of time use variables (physical activity, sedentary behaviour, and sleep)

#### 3.2.1 Physical Activity

Sixteen studies measured PA objectively using accelerometry (Table 2), whilst nine studies used subjective measures to capture PA. Subjective measures of PA included: the Athens Physical Activity Questionnaire (APAQ) (Anastasiou et al., 2018); Community Health Activity Program for Seniors (CHAMPS) questionnaire (Steinberg et al., 2015); Global Physical Activity Questionnaire (GPAQ) (Wei et al., 2021); Lifestyle Questionnaire (Marcellini et al., 2010); Physical Activity Questionnaire (PAQ) (Vance et al., 2016; Vance et al., 2005); open-ended questions about leisure time PA over past 30 days (Edwards and Loprinzi, 2017); a 7-day PA diary to capture >10 minute bouts of leisure-time PA (Lambiase et al., 2014); and a single-item questionnaire on frequency of exercise per week (only for purpose of improving health) (Yuan et al., 2020).

**Table 2.**
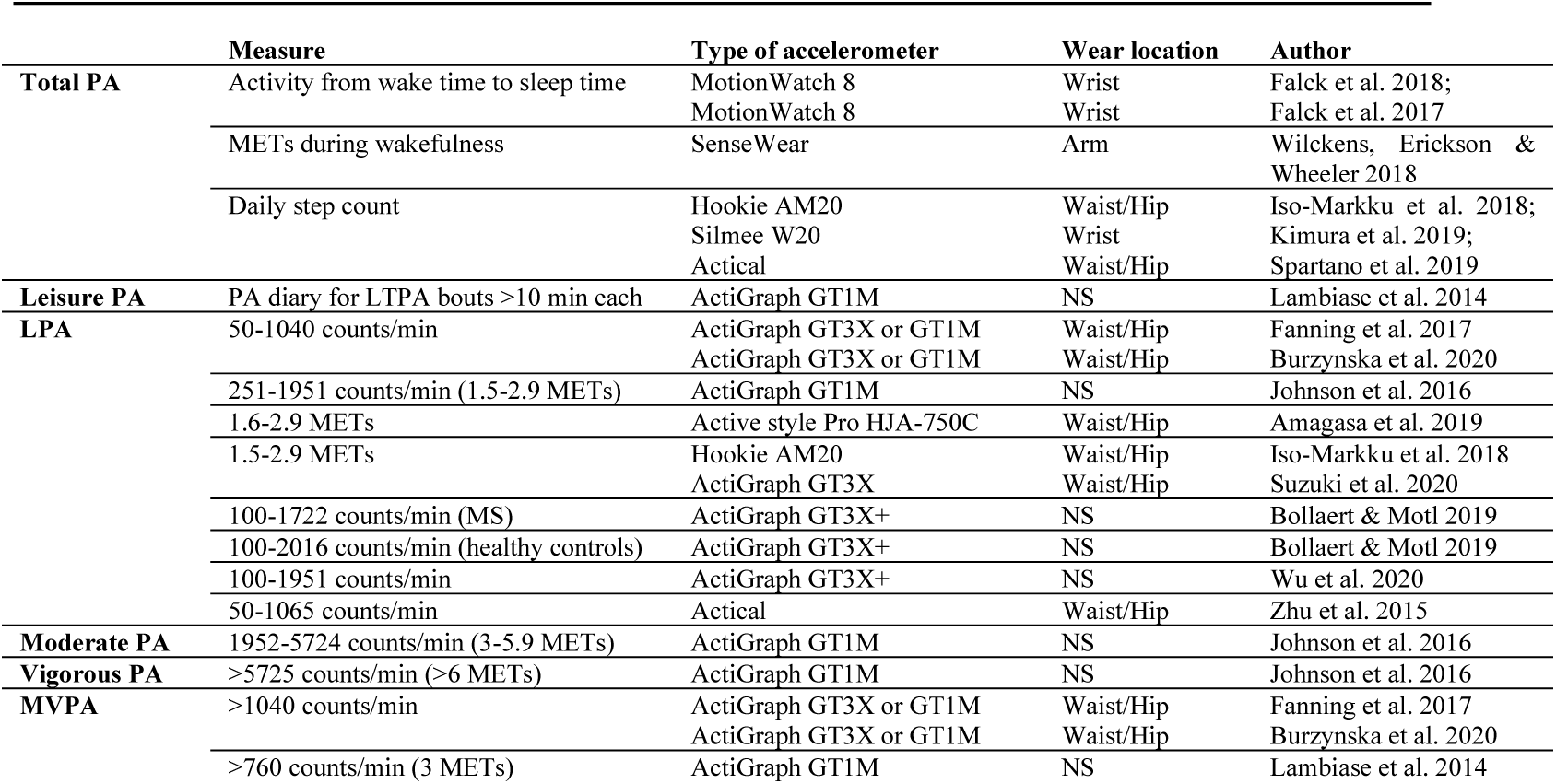
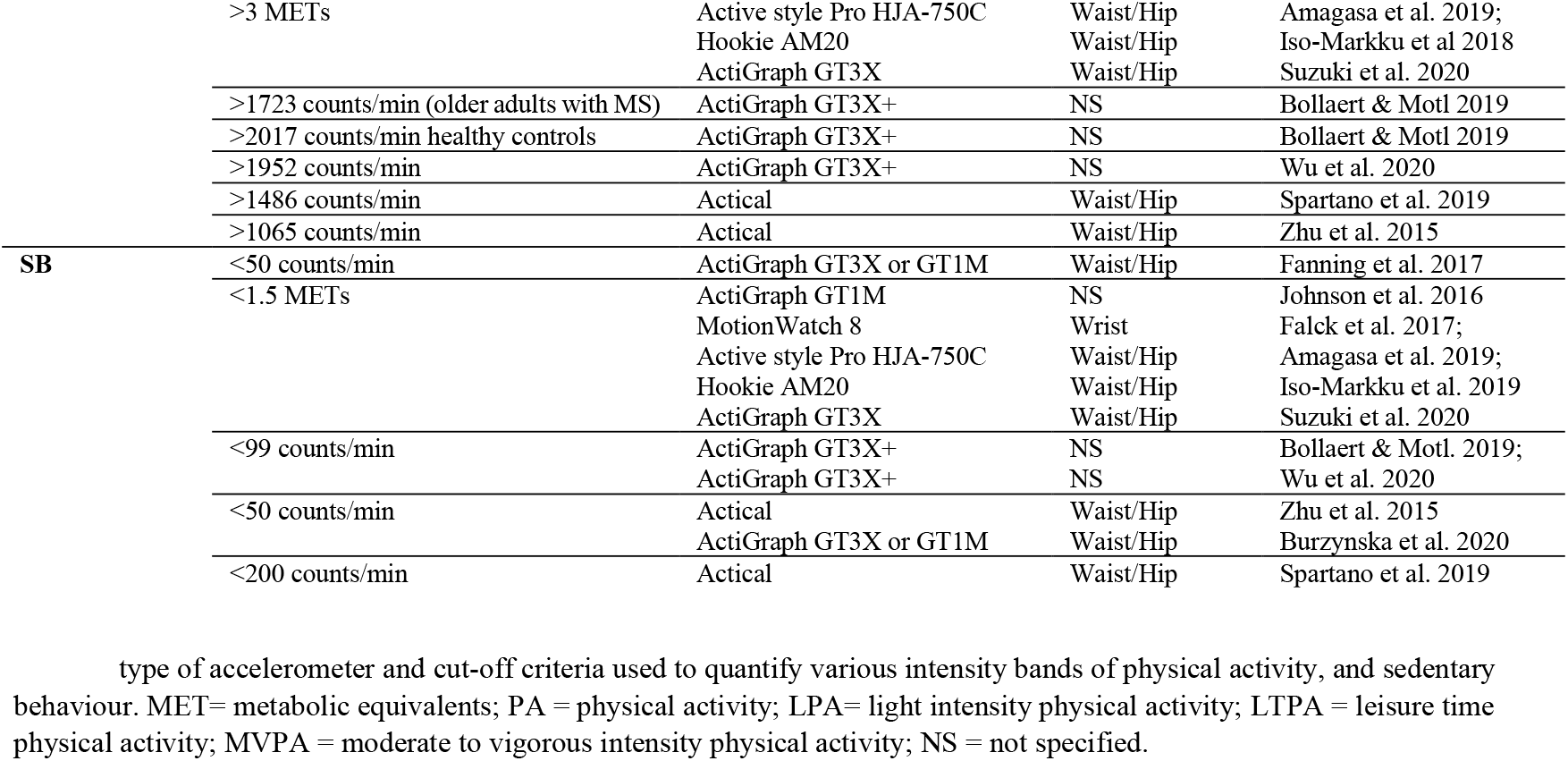
Classification of physical activity and sedentary behaviour using objective measures.

#### 3.2.2 Sedentary Behaviour

Eleven studies measured SB objectively using accelerometry (see Table 2), whilst seven studies used a subjective measure to capture SB. Subjective measures of SB included: APAQ (Anastasiou et al., 2018); GPAQ (Wei et al., 2021); CHAMPS (Steinberg et al., 2015); Lifestyle Questionnaire (Marcellini et al., 2010); PAQ (Vance et al., 2016); and two single-item questionnaires (how much time spent sitting and watching TV on a typical day over the past 30 days (Edwards and Loprinzi, 2017); how many hours spent sleeping, seated or lying down over past week (Vance et al., 2005)). In the latter study, sleep and SB were classified as the same behaviour (both grop

#### 3.2.3 Sleep

Four studies measured sleep using accelerometry. Two studies measured sleep continuously, and validated sleep onset and wake time using sleep diaries completed by participants (Lambiase et al., 2014; Wilckens et al., 2018). One study required participants to press an event marker on their wrist-worn accelerometer to mark bedtime and wake times (Falck et al., 2018). Another study measured sleep from 1800 hours to 0559 hours, with sleep start times quantified as the start of the first 20 minute block with no movement (Kimura et al., 2019).

Five studies measured sleep using self-report measures. Two captured total sleep time using language-specific versions of the Physical Activity Questionnaire (Anastasiou et al., 2018; Vance et al., 2016). Sleep quality was assessed using the Sleep Index II by Anastasiou et al. (2018), whilst Yuan et al (2020) used a 5-point Likert-type, single-question scale to capture sleep quality. The 19-item Pittsburgh Sleep Quality Index (PSQI) was used by Falck et al (2018) to capture 7 components of sleep, whilst Fanning et al (2017) used one item from the PSQI to capture average nightly sleep over the past month. Wei et al. (2021) conducted interviews to obtain values for average duration of sleep per night.

### 3.3 Associations between time-use behaviours and cognitive outcomes

The main findings of this review are displayed in Table 3 and Figure 3, whereby the results of each study are categorised into cognitive domains and summarised based on the time-use behaviours measured and relevant statistical findings. Additionally, a brief summary of the findings within each cognitive domain is provided below.

**Table 3.**
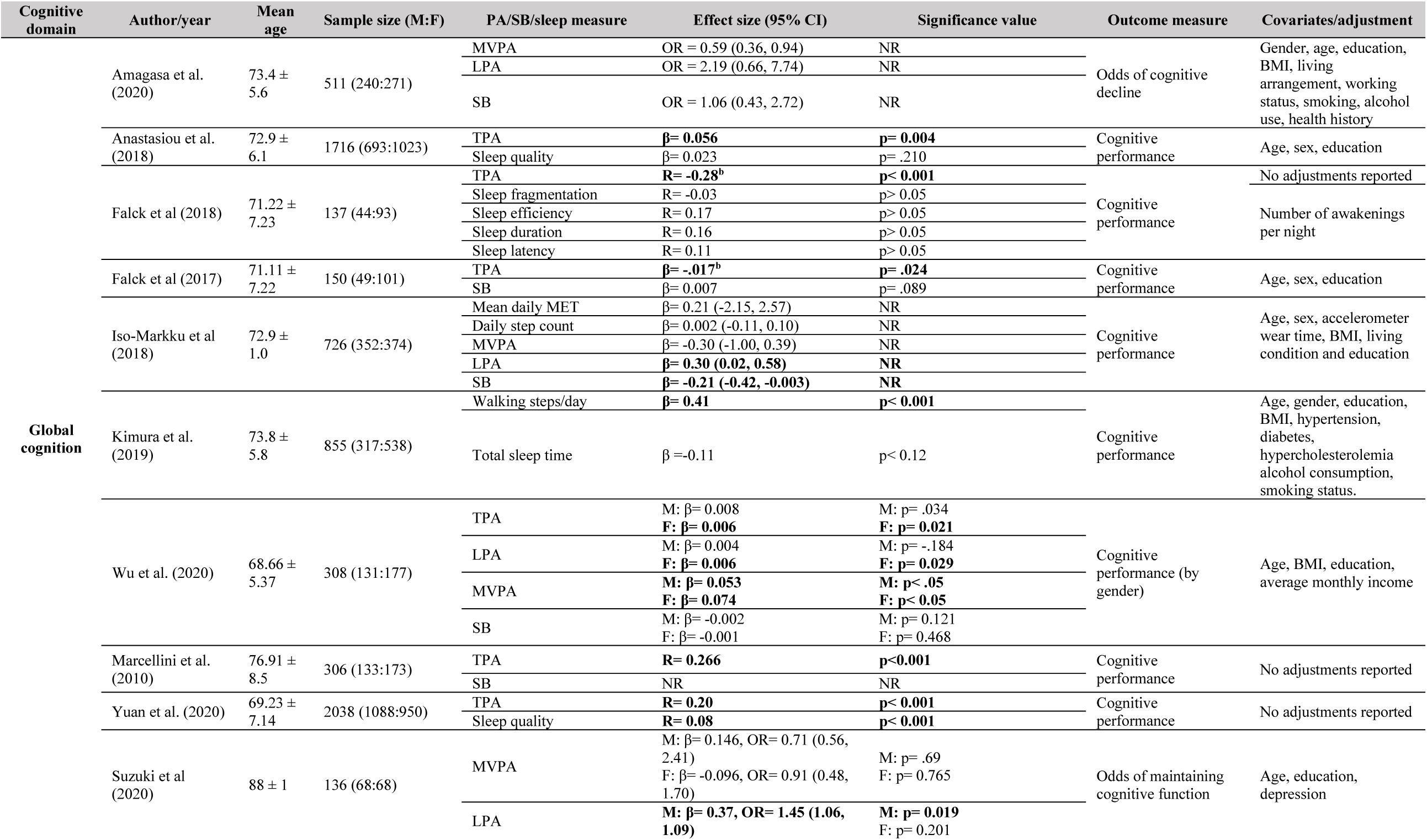
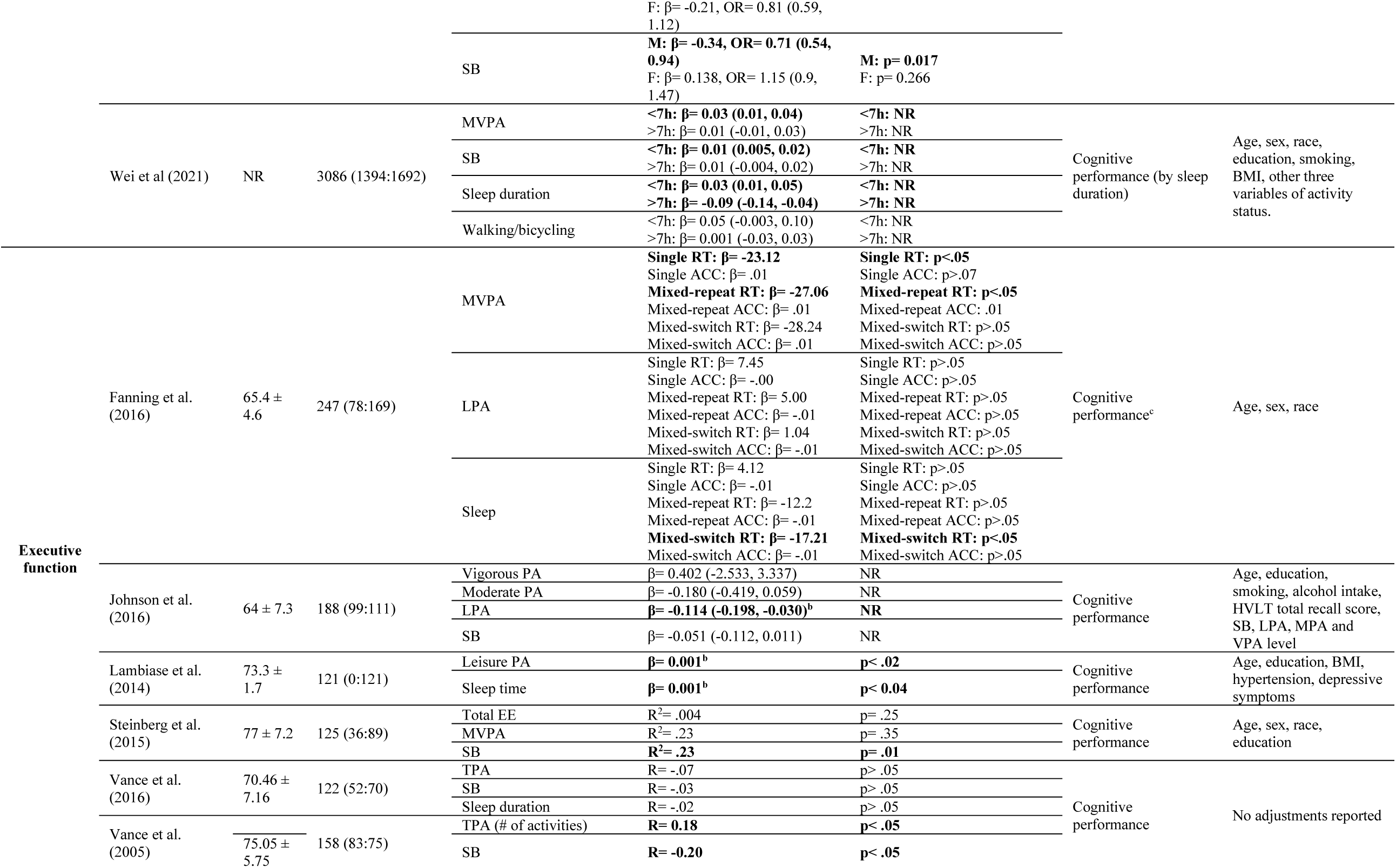
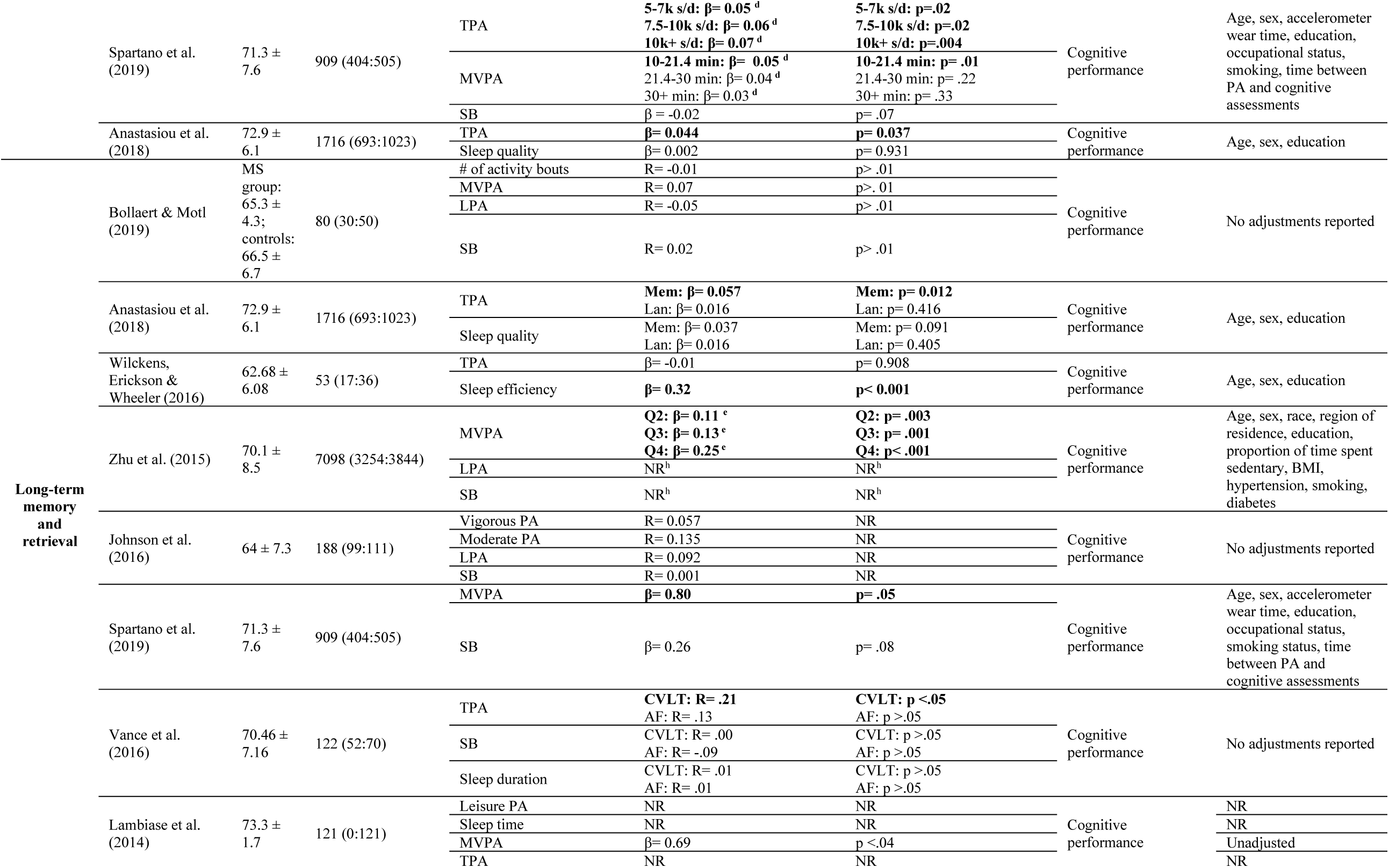
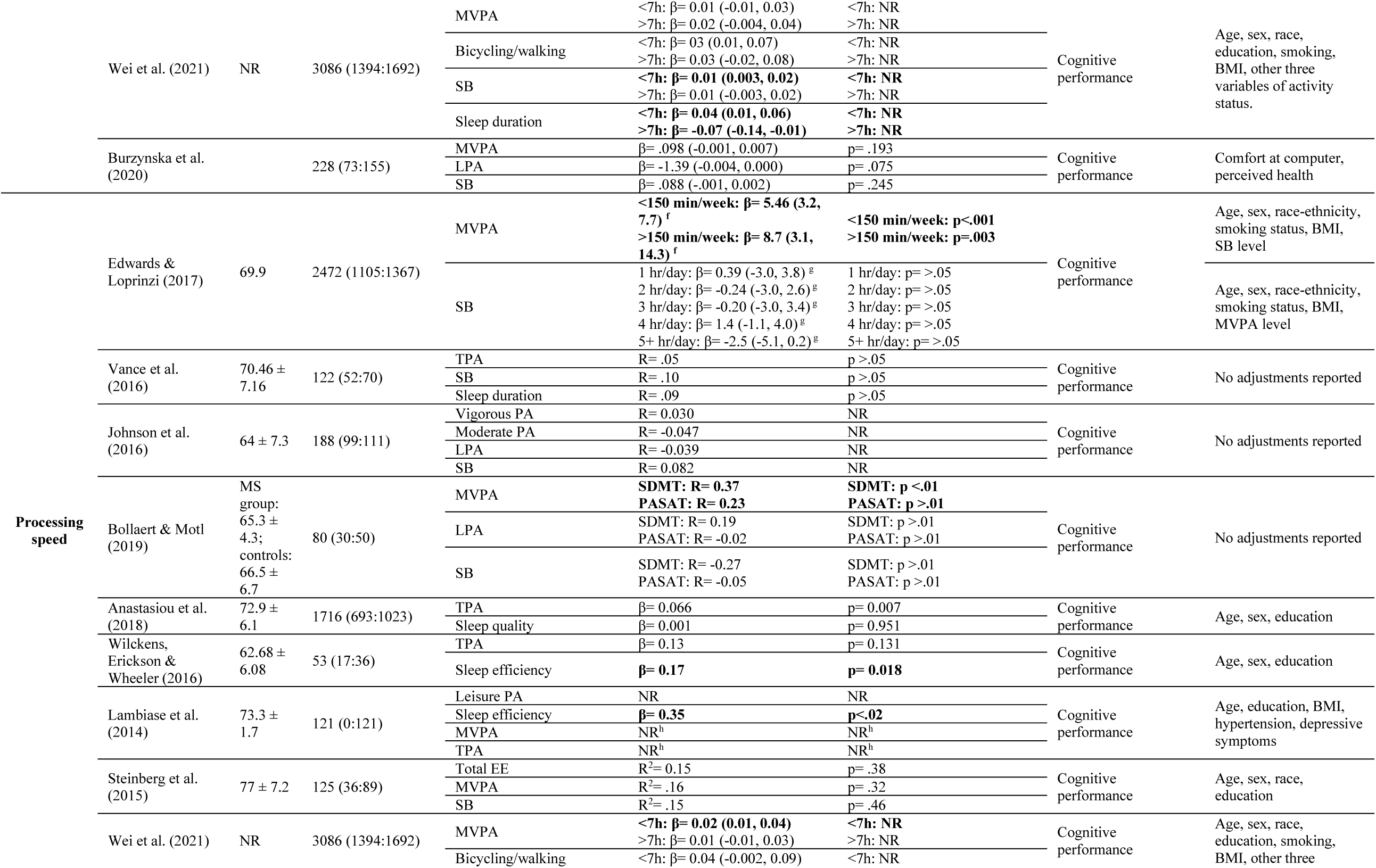
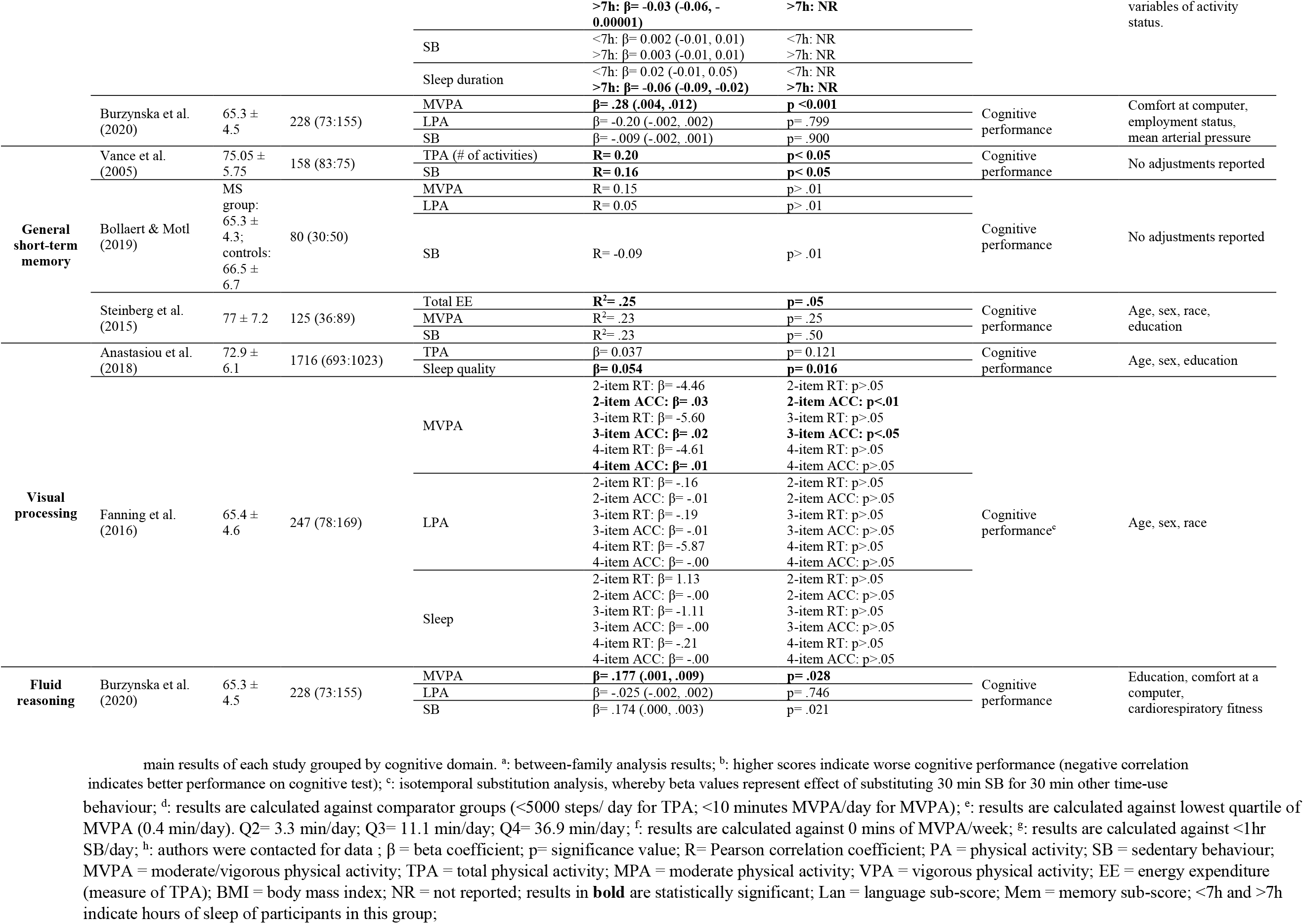
Characteristics and main results of included studies.

**Figure 3:**
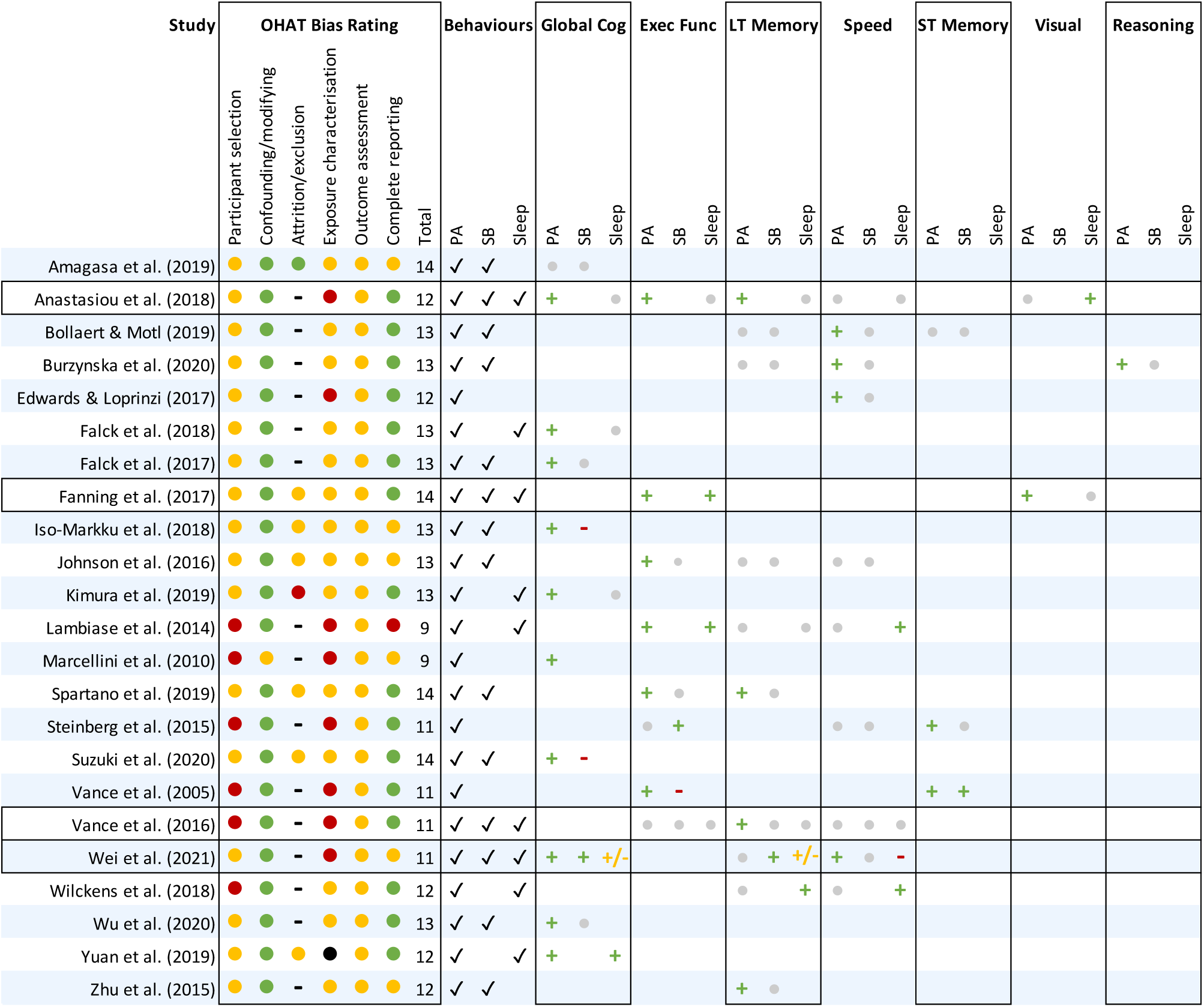
Results for the OHAT risk of bias rating tool and findings of each study by cognitive domain. OHAT= Office of Health Assessment and Translation Risk of Bias Tool, Cog= Cognition, Exec Func =Executive Function, LT= Long Term, Speed=Processing Speed, ST= Short Term, Visual= Visual Processing, Fluid= Fluid Reasoning; OHAT Scores: •=0,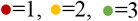; Relationships between behaviours and cognitive outcomes, **+**=positive relationship, **-**=negative relationship, **+/-**=relationship positive with some measures and negative with others, 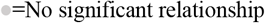, ‘-’= not reported (1 point).

#### 3.3.1 Global Cognition

The relationship between time-use behaviours and global cognition was investigated in 11 studies. The majority of these studies investigated the combination of PA and SB against global cognition (7 studies). Most frequently, spending a greater percentage of the day in PA was associated with better performance on tests of global cognition, and spending more time in SB was negatively associated with global cognition. One study by Marcellini et al. (2010) reported that lower levels of both PA and SB were associated with poorer global cognition. Several studies also reported differences in the relationship between PA intensities (MVPA and LPA) and global cognition. One study by Amagasa et al. (2019) reported that the proportion of time spent in MVPA relative to time spent in LPA or SB was significantly associated with lower odds of cognitive decline. In their study of identical twins, Iso-Markku et al. (2018) reported that LPA and SB were positively and negatively associated with global cognition, respectively, but MVPA and total PA measures were not.

Five studies investigated the association between PA and sleep with global cognition. Three studies reported a positive association between PA and global cognition but no relationships between measures of sleep and global cognition (Falck et al 2018; Anastasiou et al 2018; Yuan et al 2020). Total sleep time was found to share a negative association with global cognition when it surpassed a threshold of ∼8.35 hours per night in one study (Kimura et al 2019). Similarly, Wei et al. (2021) found that the association between sleep and global cognition was negative for older adults who engaged in >7 hours of sleep per night, but positive for those who engaged in <7 hours per night.

#### 3.3.2 Executive Function

The associations between combinations of time-use behaviours and executive function were explored in 8 studies. Five of these studies investigated the combination of PA and SB and its relationship with executive function. Overall, four studies reported that high PA/low SB combinations were positively associated with executive function, whilst only one study reported a positive association between SB and executive function (Steinberg et al. 2015). There were some discrepancies in findings of PA intensities and their associations with executive function. For example, Johnson et al. (2016) reported that LPA was positively associated with executive function, with no relationship found for SB, moderate PA or vigorous PA. Conversely, Fanning et al. (2017) reported that substituting SB with time in MVPA was associated with better executive function, whereas no association was observed when substituting SB with LPA.

Four studies investigated the associations between combinations of PA and sleep with executive function outcomes. Executive function was positively associated with time spent in PA, but not with sleep quality in a study by Anastasiou et al. (2018). In a sample of older females, greater minutes of self-reported leisure time PA was associated with *poorer* executive function, whereas shorter self-reported sleep time was associated with *better* performance on executive function (Lambiase et al., 2014). This finding contradicts Fanning et al. (2017), who found that substituting SB with time in sleep or MVPA was positively associated with executive function.

#### 3.3.3 Processing Speed

Ten studies investigated the relationship between combinations of time-use behaviours and processing speed measures. Of these, six studies reported on associations between combinations of PA and SB and processing speed. Three studies reported no association between measures of PA or SB and processing speed performance (Johnson et al., 2016; Steinberg et al., 2015; Vance et al., 2016). Two studies (Bollaert & Motl 2019; Burzynska 2020) reported positive associations between time in MVPA and processing speed, but no associations for SB or LPA.

Five studies investigated how combinations of PA and sleep were associated with processing speed. Two studies reported positive associations between PA and processing speed but no associations with sleep quality or efficiency (Anastasiou et al 2018; Wilckens, Erickson & Wheeler 2018), whereas one study reported that better sleep efficiency was associated with better processing speed (Lambiase et al., 2014). Finally, Wei et al. (2021) reported that processing speed was positively associated with time in MVPA for older adults who achieved <7hrs sleep per night, whilst SB was positively associated with processing speed in those who achieved >7hrs sleep per night.

#### 3.3.4 Long-Term Memory and Retrieval

Ten studies in this review investigated the associations between combinations of time-use behaviours and long-term memory and retrieval. Of these, seven studies investigated combinations of PA and SB. Three studies reported no associations between time spent in LPA, MVPA or SB and long-term memory and retrieval (Bollaert and Motl, 2019; Burzynska et al., 2020; Johnson et al., 2016). Two studies reported a positive association between time spent in MVPA (Zhu et al 2015; Spartano et al 2019) and total PA (Vance et al. 2016) and long-term memory and retrieval.

Five studies assessed how combinations of PA and sleep were associated with long-term memory and retrieval. One study reported no associations between PA or sleep and long-term memory (Lambiase et al 2014) whilst two studies found positive associations between TPA and long-term memory but no associations with sleep measures (Anastasiou et al. 2018; Vance et al. 2016). Finally, one study reported positive associations between sleep efficiency and long-term memory but no associations for TPA (Wilckens, Erickson & Wheeler 2018).

#### 3.3.5 General Short-Term Memory

Three studies investigated the relationship between combinations of PA and SB and general short-term memory performance. One study reported that both SB and TPA were positively associated with short-term memory (Vance et al. 2005), whilst another study reported positive associations between MVPA and short-term memory but no associations with SB (Steinberg et al 2015). Finally, one study reported no significant associations between LPA, MVPA or SB with short term memory (Bollaert & Motl).

#### 3.3.6 Visual Processing

Two studies (Anastasiou et al., 2018; Fanning et al., 2017) investigated the associations between combinations of time-use behaviours and visual processing. One study reported no relationship between PA and visual processing but a positive association with sleep quality was reported (Anastasiou et al). In another study using isotemporal substitution methods, substituting SB with MVPA was associated with better accuracy on visual processing tasks but not reaction time, whereas no associations were found for LPA or SB (Fanning et al., 2017).

#### 3.3.7 Fluid Reasoning

Only one study in this review investigated the association between combinations of time-use behaviours and fluid reasoning, reporting that both MVPA and SB were positively correlated with measures of fluid reasoning (Burzynska et al., 2020).

### 3.4 Quality and risk of bias assessment

Included studies scored between 9 and 15 (out of a possible 18) on the OHAT Risk of Bias Rating tool (Figure 3). Fifteen studies did not report on reasons for dropout/attrition, and most studies scored poorly on exposure characterisation, participant selection and outcome assessment items, indicating a probably high or definitely high risk of bias for these items. Most studies presented a probably low or definitely low risk of bias on the complete reporting item. Overall, the studies included in this review present a moderate risk of bias.

## 4. Discussion

This review aimed to summarise the existing evidence surrounding the associations between combinations of time-use behaviours (PA, SB and sleep) and cognitive function in older adults across multiple cognitive domains. Most studies investigated combinations of PA and SB, with fewer including sleep. Collectively, high PA/low SB combinations were most commonly associated with better cognitive function across domains, suggesting that spending adequate time in PA and limiting SB is beneficial for cognitive function in older age. Furthermore, while there is a relatively well-established understanding that the type and intensity of physical activity (low, moderate, vigorous) relates to cognitive outcomes, sleep and sedentary behaviour have not been looked at with the same lens. Findings suggest that, as with PA, the quality of the sleep (e.g. efficiency and continuity) and the quality of the sedentary time (e.g. cognitively passive (television) versus engaged (e.g. computer work)) are likely to be important for cognitive outcomes. Therefore, we argue that, not only is it important for future studies to measure PA, SB, and sleep (since engaging in one necessarily reduces time for the others), but also that quality should be considered. The promise of this approach includes resolving counterintuitive or seemingly contradictory study results. For example, studies suggest that long sleep may not be more beneficial for cognition than short sleep, and that there may be a middle range associated with optimal function. In addition, some studies have found that increased time in SB was associated with improved cognitive function. Broadly, developing the literature to focus on the time and quality of all three behavioural domains will be critical for future PA interventions. This research will enable consideration of the inter-dependent relationship of time use behaviours in intervention design, such that taking time from sleep versus SB to increase PA will likely lead to considerably different cognitive outcomes, as will spending SB time passively or actively, or having a long sleep of poor quality compared to a shorter sleep of better quality.

### 4.1 Combinations of physical activity and sedentary behaviour for cognitive function

The association between combinations of PA and SB with cognitive function varied considerably across cognitive domains. Most commonly, spending a greater proportion of the day in MVPA relative to other time-use behaviours was associated with better cognitive function. In addition, substituting time from SB *or* sleep to time engaging in MVPA seems to be beneficial. This finding aligns with previous studies demonstrating that engaging in MVPA is positively associated with cognitive function and various health outcomes (Rojer et al., 2021; Saunders et al., 2016). Interestingly, this review has also identified that more time spent in LPA relative to other time-use behaviours was positively associated with global cognition and executive function. Taken together, these findings support the notion that limiting SB and engaging in more PA is an effective use of time for cognitive function across several domains in older adults. Previous approaches such as the “Small Steps” program, which progressively reduced total sitting time by ∼50 mins per day following a 6-week intervention in older adults, may be an effective and sustainable approach to replacing SB with PA in older adults for cognitive benefits (Lewis et al., 2016).

Our review identified a lack of consensus about the optimal combination of time-use behaviours for cognitive function *within* cognitive domains. For example, within the executive function domain, one study reported a positive association between time spent in LPA and executive function (Johnson et al., 2016), whilst another reported no association (Fanning et al., 2017). One potential reason for the discrepancy may be between-study differences in cut-points used to classify LPA from the raw accelerometry data (see Table 2). Johnson et al. (2016) classified LPA as activity which fell between 251 and 1951 counts/minute, whereas Fanning et al. (2017) used a lower cut point of 50 counts/minute, and an upper cut point of 1040 counts/minute to classify LPA. The LPA cut points used by Johnson et al. (2016) may have classified some MVPA as LPA, given the high upper threshold of 1951 counts/minute, and similarly, Fanning et al. (2017) may have captured sedentary behaviours as LPA given the considerably lower threshold of 50 counts/minute. Given the considerable heterogeneity in research approaches, meta-analysis of results was not feasible, thus limiting conclusions about optimal combinations of MVPA, LPA and SB for cognitive function.

### 4.2 Combinations of physical activity and sleep for cognitive function

Several notable commonalities between studies were identified for sleep and PA combinations. First, findings alluded to a window of optimal sleep duration for enhancing cognitive function in older adults. Studies reported negative associations of achieving >7 hours (Wei et al., 2021) and >8.25 hours (Kimura et al., 2019) of sleep per night, respectively, with cognitive function. Similarly, Lambiase et al. (2014) reported that participants with shorter self-reported sleep performed better in tests of executive function. The findings of this review align with previous studies suggesting that duration of sleep above or below the recommended range (<6h, >8h) in older adults is associated with poorer cognitive performance and greater risk of dementia (Benito-León et al., 2009; Benito-León et al., 2014; Lo et al., 2016; Spira et al., 2017; Westwood et al., 2017).

Sleep is multi-faceted, and measures of sleep duration alone do not capture the “whole picture” of sleep experiences (Falck et al., 2021). Sleep efficiency, traditionally defined as the ratio of actual sleep time to time in bed (Reed and Sacco, 2016), serves as an important measure of the sleep experience in older adults who typically have more fragile sleep patterns than younger adults (Mander et al., 2017). Older adults with longer sleep durations often report poorer sleep quality and increased sleep fragmentation, and the latter has been proposed as a potential mechanism for the relationship between long sleep duration and increased mortality (Grandner and Drummond, 2007). In this review we found that measures of sleep efficiency were frequently associated with better cognitive function independently of PA (Falck et al., 2018; Lambiase et al., 2014; Wilckens et al., 2018). Previous studies have also demonstrated that sleep disruption is associated with poorer cognitive function in older adults with (Naismith et al., 2010) and without (Miyata et al., 2013) mild cognitive impairment, particularly for measures of executive function. In summary, given the complex nature of sleep it is important to investigate sleep quality measures in addition to time spent in sleep to gain a comprehensive understanding of optimal sleep for cognitive function in older adults (Falck et al., 2021).

Importantly, several studies in this review discussed the interactions between sleep and PA for cognitive function in older adults. One study reported that greater PA levels modified the relationship between poor sleep efficiency and processing speed in participants with low activity levels (Lambiase et al., 2014), whilst another reported that reallocating sleep time to any form of PA was associated with better global cognition in those who slept >7h/night (Wei et al. 2021). Taken together, these findings suggest that engaging in PA may attenuate the negative effects of poor sleep on cognitive function in older adults. The importance of interactions between PA and sleep are becoming increasingly recognised. It is understood that increasing PA engagement can improve sleep quality in older adults (Gubelmann et al., 2018). Furthermore, a previous study of middle-aged women by Patel et al. (2006) identified that the likelihood of engaging in long sleep durations was associated with low levels of exercise, and conversely, those engaging in higher levels of exercise were less likely to engage in long sleep. It is recommended that future studies investigate the interactions between PA and sleep for cognitive function, and incorporate measures of sleep in addition to sleep duration, to gain a holistic understanding of the best practice sleep and PA patterns for cognitive function in older adults.

### 4.3 Types of sedentary behaviour are differentially related to cognitive function

A resounding finding of studies in this review was that high SB/low PA was negatively associated with cognitive function outcomes. Yet, four studies reported that *more* time spent in SB was *positively* associated with cognitive function (Burzynska et al., 2020; Marcellini et al., 2010; Vance et al., 2005; Wei et al., 2021). This may be due to several factors. First, previous research has suggested that different types of SB may have diverging effects on cognitive function. A recent review reported that total SB and watching TV were most frequently associated with unfavourable health outcomes, whereas computer use was associated with positive outcomes in cognitive function (Ross et al., 2020). It may be that more cognitively demanding sedentary leisure activities (e.g., those subjectively measured in the study by Marcellini et al. (2010) such as reading or playing cards) are positively associated with cognitive function, whereas cognitively passive activities such as sitting and watching TV are not. Future research should aim to combine objectively measured (e.g. accelerometry) SB with 24-hour recall measures, to obtain further information on the context of SB and how this may influence its relationship with cognitive function (Burzynska et al., 2020). Building an understanding of the relationships between different types of SB with cognitive function is an important step in providing informed SB guidelines for cognitive function in older adult populations.

Notably, the study by Vance et al. (2005) which reported a positive association between time spent in SB and cognitive function used a single self-report question (*how many hours per day do you spend sitting, sleeping or lying down?)* as a measure of SB, but did not measure sleep as a separate time-use behaviour. Therefore, in this study, sleep was considered a SB. This approach is a limitation as there are likely to be considerable differences between the associations of sleep and SB with cognitive function. Including time spent in sleep as a SB limits the ability to determine whether sleep or SB is driving the beneficial association with cognitive function, and this approach should be avoided in future studies.

### 4.4 Considering time use as a three-part composition

The importance of considering multiple time-use behaviours in the same statistical model is becoming increasingly recognised in health research, driving the shift to a ‘time-use epidemiology’ paradigm (Pedišić et al., 2017). Considering PA, SB and sleep as separate or independent behaviours limits the application of conclusions to holistic 24-hour interventions and movement guidelines. Only two studies meeting review inclusion criteria assessed all three time-use behaviours together in one model (Fanning et al., 2017; Wei et al., 2021). Fanning et al. (2017) reported that substituting time spent in SB with MVPA or sleep was associated with better cognitive outcomes; however, substituting SB with LPA was not associated with any beneficial cognitive outcomes. Wei et al. (2021) also reported that for participants who achieved <7h of sleep per night, substituting SB with any form of PA was associated with better global cognition, whereas in participants who achieved >7h of sleep per night, substituting sleep with PA or SB was associated with better global cognition. Considering all three time-use behaviours in one model allows the compensatory follow-on ripple effects of changing one behaviour (e.g., increasing PA) across the daily 24-hour composition (e.g., decreasing sleep and/or SB) to be considered. It should be noted that there were several other studies in this review which did investigate three time-use behaviours, but they were not included in the same model.

### 4.5 Study limitations

A few study limitations should be considered. The small number of studies within each cognitive domain limits the strength of conclusions that can be made about the best combinations of time-use behaviours for individual cognitive domains. As there was considerable variability in the methods used to measure PA, sleep and SB between studies in this review, we were unable to conduct a meta-analysis to determine the overall strength of the relationship between combinations of time-use behaviours and cognitive function. Further, no grey literature or unpublished material was included in this review. The use of compositional data analysis approaches, including isotemporal substitution methods, is becoming increasingly popular and so exclusion of unpublished works may have missed relevant findings.

### 4.6 Conclusion

This review identified that spending more time in PA and limiting SB was associated with better cognitive outcomes in older adults. Higher proportions of MVPA in the day were most frequently associated with better cognitive function, and some evidence highlighted the benefits of spending higher proportions of the day in LPA for global cognition and executive function domains only. Spending higher proportions of the day in SB was negatively associated with cognitive function in most studies, although there was evidence to suggest certain types of SB may be positively associated with cognitive function. Furthermore, several studies in this review support the idea that sleep duration shares an inverted U-shaped relationship with cognitive function, such that sleep durations above or below the ‘normal range’ (∼7-9 hours) are negatively associated. Alternative measures to capture sleep quality characteristics such as sleep efficiency are positively associated with cognitive function, highlighting the importance of using multiple measures of sleep to more fully understand optimal sleep patterns for cognitive function in older adults. Currently only a small number of studies have investigated time-use behaviours as parts of a 24-hour time-use composition against cognitive function. To more fully understand and exploit the opportunities of time-use interventions for the prevention or delay of many non-communicable diseases, the 24-hour time-use approach is crucial for future research. Additionally, there is considerable heterogeneity in methodological and statistical approaches used in this field thus far. A more standardised approach to capturing PA, SB and sleep in older adults is required.

## Data Availability

Data are available via request to the authors.

## 5. Acknowledgements

This work was supported by the National Health and Medical Research Council (NHMRC) [grant number 1171313]. M.L.M. is supported by a Dementia Australia Research Foundation PhD scholarship. D.D. is supported by an NHMRC Early Career Fellowship (GNT1162166) and the National Heart Foundation of Australia (1020840). M.R.G. is supported by an Australian Research Council (ARC) fellowship (DE200100575).

