## Supplementary File A and B for "How are combinations of physical activity, sedentary behaviour and sleep related to cognitive function in older adults? A systematic review"

**Supplementary Material 1**

**Search Strategy**

1. Exercise/

2. Sports/

3. (Physical* activ* or sport* or garden* or golf* or lawn bowl* or yoga or tai chi or meditat* or walk* or jog* or run* or bicycl* or gym or physical train* or aerobic exercis* or aerobic train*).mp. [mp=title, abstract, original title, name of substance word, subject heading word, floating sub-heading word, keyword heading word, organism supplementary concept word, protocol supplementary concept word, rare disease supplementary concept word, unique identifier, synonyms]

4. 1 or 2 or 3

5. Sedentary Behavior/

6. sedentar*.mp. [mp=title, abstract, original title, name of substance word, subject heading word, floating sub-heading word, keyword heading word, organism supplementary concept word, protocol supplementary concept word, rare disease supplementary concept word, unique identifier, synonyms]

7. (chair time or desk time or car time or bus time or indoor time or screen time or computer time).mp. [mp=title, abstract, original title, name of substance word, subject heading word, floating sub-heading word, keyword heading word, organism supplementary concept word, protocol supplementary concept word, rare disease supplementary concept word, unique identifier, synonyms]

8. low energy expenditure.mp. [mp=title, abstract, original title, name of substance word, subject heading word, floating sub-heading word, keyword heading word, organism supplementary concept word, protocol supplementary concept word, rare disease supplementary concept word, unique identifier, synonyms]

9. (computer game* or video game* or television or tv).mp. [mp=title, abstract, original title, name of substance word, subject heading word, floating sub-heading word, keyword heading word, organism supplementary concept word, protocol supplementary concept word, rare disease supplementary concept word, unique identifier, synonyms]

10. (screen based entertainment or screen-based entertainment).mp. [mp=title, abstract, original title, name of substance word, subject heading word, floating sub-heading word, keyword heading word, organism supplementary concept word, protocol supplementary concept word, rare disease supplementary concept word, unique identifier, synonyms]

11. bed rest.mp. [mp=title, abstract, original title, name of substance word, subject heading word, floating sub-heading word, keyword heading word, organism supplementary concept word, protocol supplementary concept word, rare disease supplementary concept word, unique identifier, synonyms]

12. sitting.mp. [mp=title, abstract, original title, name of substance word, subject heading word, floating sub-heading word, keyword heading word, organism supplementary concept word, protocol supplementary concept word, rare disease supplementary concept word, unique identifier, synonyms]

13. physical* inactiv*.mp. [mp=title, abstract, original title, name of substance word, subject heading word, floating sub-heading word, keyword heading word, organism supplementary concept word, protocol supplementary concept word, rare disease supplementary concept word, unique identifier, synonyms]

14. 5 or 6 or 7 or 8 or 9 or 10 or 11 or 12 or 13

15. Sleep/

16. Sleep Deprivation/

17. sleep* durati*.mp. [mp=title, abstract, original title, name of substance word, subject heading word, floating sub-heading word, keyword heading word, organism supplementary concept word, protocol supplementary concept word, rare disease supplementary concept word, unique identifier, synonyms]

18. 15 or 16 or 17

19. 4 and 14

20. 4 and 18

21. 14 and 18

22. 4 and 14 and 18

23. 19 or 20 or 21 or 22

24. Cognition/

25. (Cogniti* or attention or awareness or psychomotor perform* or comprehensi* or consciousness or executive func* or learn* or problem solv* or planning or decision? making or memory or perception or inhibit* or object naming or word finding or fluency or visuospatial or processing speed).mp. [mp=title, abstract, original title, name of substance word, subject heading word, floating sub-heading word, keyword heading word, organism supplementary concept word, protocol supplementary concept word, rare disease supplementary concept word, unique identifier, synonyms]

26. 24 or 25

27. 23 and 26

28. limit 27 to ("middle aged (45 plus years)" and english)

29. limit 28 to (("middle aged (45 plus years)" or "all aged (65 and over)") and english)

**Supplementary Material 2**

OHAT Risk of Bias Rating Tool items:

1. *Participant selection:* Did selection of study participants result in appropriate comparison groups?
2. *Confounding/modifying:* Did the study design or analysis account for important or confounding and modifying variables?
3. *Attrition/exclusion:* Were outcome data complete without attrition or exclusion from analysis?
4. *Exposure characterisation:* Can we be confident in the exposure characterisation?
5. *Outcome assessment:* Can we be confident in the outcome assessment?
6. *Complete reporting:* Were all outcome measures reported?

OHAT Risk Of Bias Rating Tool: full results and ratings

|  | Amagasa et al. (2019) | Anastasiou et al. (2018) | Bollaert & Motl (2019) | Burzynska et al. (2020) | Edwards & Loprinzi (2017) | Falck et al. (2018) | Falck et al. (2017) | Fanning et al. (2017) | Iso-Markku et al. (2018) | Johnson et al. (2016) | Kimura et al. (2019) | Lambiase et al. (2014) | Marcellini et al. (2010) | Spartano et al. (2019) | Steinberg et al. (2015) | Suzuki et al. (2020) | Vance et al. (2005) | Vance et al. (2016) | Wei et al. (2021) | Wilckens et al. (2018) | Wu et al. (2020) | Yuan et al. (2020) | Zhu et al. (2015) |
| --- | --- | --- | --- | --- | --- | --- | --- | --- | --- | --- | --- | --- | --- | --- | --- | --- | --- | --- | --- | --- | --- | --- | --- |
| Did selection of study participants result in appropriate comparison groups? | 2 | 2 | 2 | 2 | 2 | 2 | 2 | 2 | 2 | 2 | 2 | 1 | 1 | 2 | 1 | 2 | 1 | 1 | 2 | 1 | 2 | 2 | 2 |
| Did the study design or analysis account for important confounding and modifying variables? | 3 | 3 | 3 | 3 | 3 | 3 | 3 | 3 | 3 | 3 | 3 | 3 | 2 | 3 | 3 | 3 | 3 | 3 | 3 | 3 | 3 | 3 | 3 |
| Were outcome data complete without attrition or exclusion from analysis? | 3 | NR | NR | NR | NR | NR | NR | 2 | 2 | 2 | 1 | NR | NR | 2 | NR | 2 | NR | NR | NR | NR | NR | 2 | NR |
| Can we be confident in the exposure characterisation? | 2 | 1 | 2 | 2 | 1 | 2 | 2 | 2 | 2 | 2 | 2 | 1 | 1 | 2 | 1 | 2 | 1 | 1 | 1 | 2 | 2 | 0 | 2 |
| Can we be confident in the outcome assessment? | 2 | 2 | 2 | 2 | 2 | 2 | 2 | 2 | 2 | 2 | 2 | 2 | 2 | 2 | 2 | 2 | 2 | 2 | 2 | 2 | 2 | 2 | 2 |
| Were all outcome measures reported? | 2 | 3 | 3 | 3 | 3 | 3 | 3 | 3 | 2 | 2 | 3 | 1 | 2 | 3 | 3 | 3 | 3 | 3 | 2 | 3 | 3 | 3 | 2 |
| **TOTAL SCORE (/18)** | **14** | **12** | **13** | **13** | **12** | **13** | **13** | **14** | **13** | **13** | **13** | **9** | **9** | **14** | **11** | **14** | **11** | **11** | **11** | **12** | **13** | **12** | **12** |
